# Transitional care needs of persons with dementia and their care partners: a secondary analysis using Transitions Theory

**DOI:** 10.64898/2026.06.24.26356479

**Authors:** Cheng Li, Wanyi Gwenneth Wang, Katherine LoVerde, Komal Patel Murali

**Affiliations:** Rory Meyers College of Nursing, New York University, New York, New York, United States of America; College of Arts & Science, New York University, New York, New York, United States of America; NYU Langone Health, New York, New York, United States of America

## Abstract

Persons with dementia (PWD) and their care partners experience multidimensional care transitions across the illness trajectory. Care transitions interventions often center the experience of transfer between healthcare settings and depend on specialized services, which limits incorporation into standard routine care. This study examined the transitional care needs of PWD and care partners using Transitions Theory to inform transitional care interventions, especially for hospice care transitions. This is a secondary qualitative directed content analysis. Data were coded using Transitions Theory concepts, analyzed using framework analysis, including data familiarization, framework identification, indexing, charting, and interpretation. Participants (n=20) with an average age 53 were mostly female children care partners (80%, n=16) of PWD identifying as Black, Hispanic/Latino, Asian, or White. Five themes were identified: (1) “They change and you taking the role now of the parent”: Multiplicity and complexity of transitions, identifying overlapping developmental, health-illness, situational, and organizational transitions; (2) Transitions occur in unanticipated ways, reflecting variable awareness and unpredictable illness trajectories; (3) Challenging transition conditions, including personal, community, and systemic factors; (4) Challenging decision making in transitions, highlighting care partners’ uncertainty and moral distress making ongoing decisions; and (5) Critical supports for healthy transitions, identifying guidance, care network, psychosocial well-being, and community resources as critical for transitions. Transitional care in dementia is characterized by multiplicity and unpredictability of transitions. When tailored transitional care guidance is lacking, care partners must take on significant responsibility for finding information and making decisions. Transitional care interventions that integrate anticipatory guidance, care planning facilitation, psychosocial support, and community linkage are critical to addressing these needs.

## Introduction

Dementia is a progressive condition that profoundly affects individuals with the illness and those who support them. As cognitive, functional, and communicative abilities decline over time, persons with dementia (PWD) increasingly rely on care partners to navigate complex healthcare systems. These experiences are accompanied by a series of transitions that occur across the dementia trajectory, both PWD and care partners experience these transitions that are often complex, unanticipated, and unpredictable. Effective management of care transitions is critical to maintaining continuity of care, supporting decision-making, and promoting quality of life for both PWD and their care partners [1].

Transitions Theory [2, 3] provides a framework to understand the multidimensional experiences of PWD and care partners during care transitions. In the theory, transitions are dynamic and ongoing processes, triggered by changes that necessitate adaptation in roles, responsibilities, and daily routines; influenced by personal, relational, and contextual factors. The theory conceptualizes four types of transitions and emphasizes the properties of transition experiences, transition conditions, and patterns of response [2]. Applying Transitions Theory allows for a systematic exploration of the evolving transitional care needs of PWD and their care partners across the dementia trajectory, forming a conceptual basis for developing practical interventions that are integrated into routine care.

Research on dementia transitions has primarily focused on care setting changes, such as moving from home to hospital, returning home, or entering residential care (Saragosa et al., 2022). Few on the broader illness trajectory from before diagnosis to end-of-life care [4, 5], fewer on detailed transitions such as becoming the primary decision-maker and adjusting to increased responsibility [6].

Transitional care is critical for comprehensive dementia care, given the progressive and unpredictable nature of the illness [7]. Current interventions for transitional care in dementia [1] often rely on counseling or care management, which may involve regular assessment and follow-up, requiring healthcare professionals to deliver complex care guidance and supportive care. However, these models can be optimized through the incorporation of transitional care considerations for PWD and care partners. Consequently, there is a need to identify broader transitional care needs that can serve as a foundation for further individualized assessment.

This study represents a qualitative secondary analysis of data from a parent study exploring the views and experiences of dementia partners receiving home healthcare at the end of life. The purpose of this analysis is to identify the transitional care needs of PWD and their care partners through a theory-driven approach using Transitions Theory. By applying this lens, the study seeks to provide a foundation for assessment to inform transitional care into routine healthcare practice.

## Methods

This secondary analysis used de-identified qualitative data from a parent study examining hospice and end-of-life care transitions in home healthcare. Participants are 20 racially and ethnically diverse care partners of PWD identified through ICD-10 diagnosis codes, with PWD meeting care partner-reported Quick Dementia Rating Scale [8] criteria for moderate to severe dementia. Recruitment, screening, and semi-structured interviews occurred from October 2023 to January 2024 at NYU and VNS Health.

The study was approved by the institutional review boards of NYU Langone Medical Center and VNS Health. Data were collected and initially coded during the parent study. For this analysis, de-identified data was accessed on January 23, 2026. After initial rounds of familiarization with the data and parent study codebook, transcripts were coded using Transitions Theory concepts with a framework analysis approach [9, 10]. The analysis steps are:(1) data familiarization, (2) framework identification, (3) data to framework indexing, (4) indexed data summary charting, and (5) mapping and interpretation of patterns found in the charts. This approach allowed exploration of themes related to transitional care needs of dementia care partners receiving home healthcare in later stages of illness.

### Reflexivity & Rigor

Rigor in this study was established through strategies that enhance reflexivity, credibility, authenticity, integrity [11]. Reflexivity was an ongoing process throughout the study. The first author is a nursing professional and a fifth-year family caregiver to two individuals with Alzheimer’s disease. This dual role informed the conceptualization of the study and sensitivity to the complexities of dementia care. Professional training in nursing contributed an understanding of the clinical realities of dementia progression, care coordination, and healthcare systems. While familial caregiving experience offered insight into the relational and practical dimensions of navigating care over time. Team discussions were used to examine how researchers’ backgrounds shape the research process, thereby enhancing analytic transparency and rigor.

Credibility was supported through peer debriefing with the research team to critically examine analytic decisions. Authenticity was maintained by ensuring a fair and balanced representation of participants’ experiences, capturing diverse perspectives across care partners. The use of rich, verbatim quotations provides transparency and allow participants’ voices to be directly reflected in the findings. Integrity was ensured through systematic and transparent data analysis procedures. A clear and detailed documentation of the research process was performed, including methodological decisions, coding development, and analytic memos. These strategies support the trustworthiness and rigor of the study.

### Framework analysis

As an initial step in data familiarization, the coded data from parent study were reviewed applying Transitions Theory. Using this theory to operationalize the analytical framework, the preliminary codes were subsequently delinked, and the original transcripts were re-examined line by line. To ensure that the data reflected a broader dementia illness trajectory and captured multiple types of transitions, the interview guide was revisited. It was confirmed that in addition to hospice transitions, discussions were expanded to include care partners’ overall experiences of dementia care. Key ideas related to care partners’ experiences were documented through notetaking.

Transitions Theory (Meleis et al., 2000) was then examined in depth and selected as the framework for analysis. While the original conceptual model was considered structurally supportive, several refinements were made to better align it with the data. Specifically, the component “transition time span” was removed, and subcomponents within “process indicators,” and “outcome indicators” were collapsed. The refined framework was tested against one-quarter of the dataset, after which formal line-by-line coding was initiated using the retained framework concepts as codes.

This coding phase functioned as an indexing process. During this stage, the concepts of “awareness” and “engagement” were merged; ‘single’ ‘sequential’ ‘unrelated’ under patterns were removed to enhance analytic fit and coherence with the data. Through systematic charting, earlier analytic decisions were revisited, ensuring appropriate levels of data abstraction and confirming the adequacy of the refined framework. The results of charting, along with the final stage of mapping and interpretation, are presented in Table 2.

**Table 1.** Characteristics of Care Partners and PWD.

| Characteristic | Care Partners (n = 20) | PWD (n = 21) |
| --- | --- | --- |
| **Demographics** |  |  |
| **Age (Mean ± SD)** | 52.7 ± 16.2 | 81.2 ± 8.57 |
| **Age Range** | 22-80 | 65-95 |
| **Gender** | N (%) | N (%) |
| **Woman** | 16 (80%) | 18 (85.7%) |
| **Man** | 4 (20%) | 3 (14.3%) |
| **Race** | N | N |
| **Black/African American** | 5 (25%) | 5 (23.8%) |
| **Non-Hispanic White** | 2 (10%) | 2 (9.5%) |
| **Hispanic White** | 3 (15%) | 4 (19%) |
| **Asian** | 7 (35%) | 8 (38.1%) |
| **Bi- or Multiracial** | 3 (15) | 2 (9.5%) |
| **Ethnicity** | N (%) | N (%) |
| **Hispanic** | 5 (25%) | 5 (23.8%) |
| **Non-Hispanic** | 11 (55%) | 10 (47.6%) |
| **Other (e.g., Chinese, Indian)** | 4 (20%) | 6 (28.6%) |
| **Education / Insurance** | N(%) | N (%) |
| **High School/GED** | 4 (20%) | — |
| **Some college** | 4 (20%) | — |
| **Bachelor's Degree** | 7 (35%) | — |
| **Master's Degree** | 3 (15%) | — |
| **Professional/Doctorate** | 2 (10%) | — |
| **Medicare** | — | 4 (19%) |
| **Medicaid** | — | 2 (9%) |
| **Dual Eligible** | — | 14 (67%) |
| **Private/Commercial** | — | 1 (5%) |
| **Relationship to PWD** | N (%) | — |
| **Spouse** | 3 (15%) | — |
| **Adult Child** | 12 (60%) | — |
| **Niece/Nephew** | 1 (5%) | — |
| **Grandchild** | 2 (10%) | — |
| **Sibling** | 2 (10%) | — |
| **Years as Caregiver (n = 10)** | 8.4 ± 7.2 | — |
| **Clinical Characteristics (PLWD) N(%)** |  |  |
| **Length of Home Care** | — | <1 year: 7 (33%)<br>1–5 years: 8 (38%)<br>6–10 years: 5 (24%)<br>>20 years: 1 (5%) |
| **Diagnosis Type** | — | Alzheimer's Disease: 9 (43%)<br>Dementia NOS: 6 (29%)<br>Early-Onset Dementia: 1 (5%)<br>Lewy Body Dementia: 1 (5%)<br>Vascular Dementia: 1 (5%)<br>Parkinson's + Dementia: 2 (10%) |

**Table 2.**
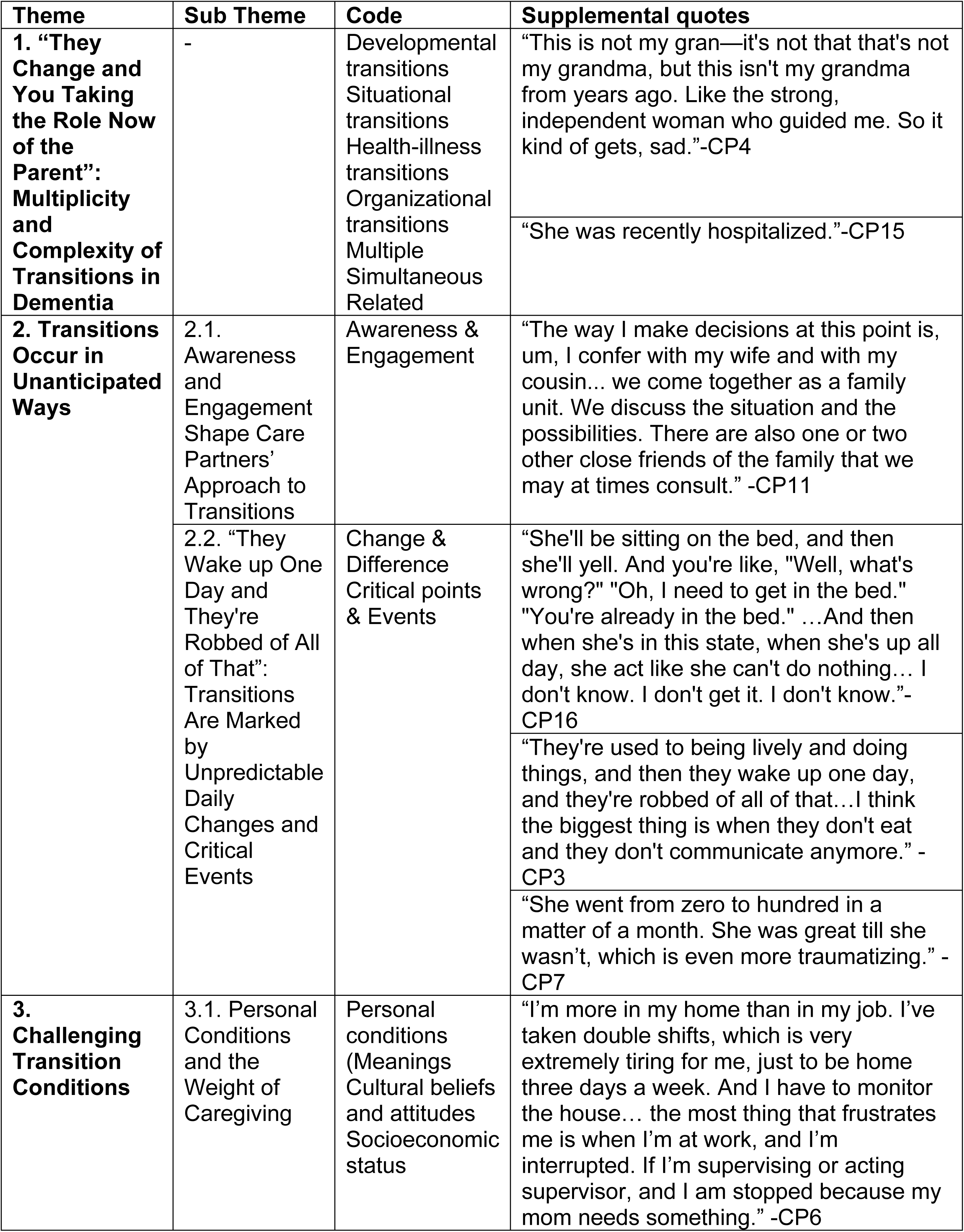
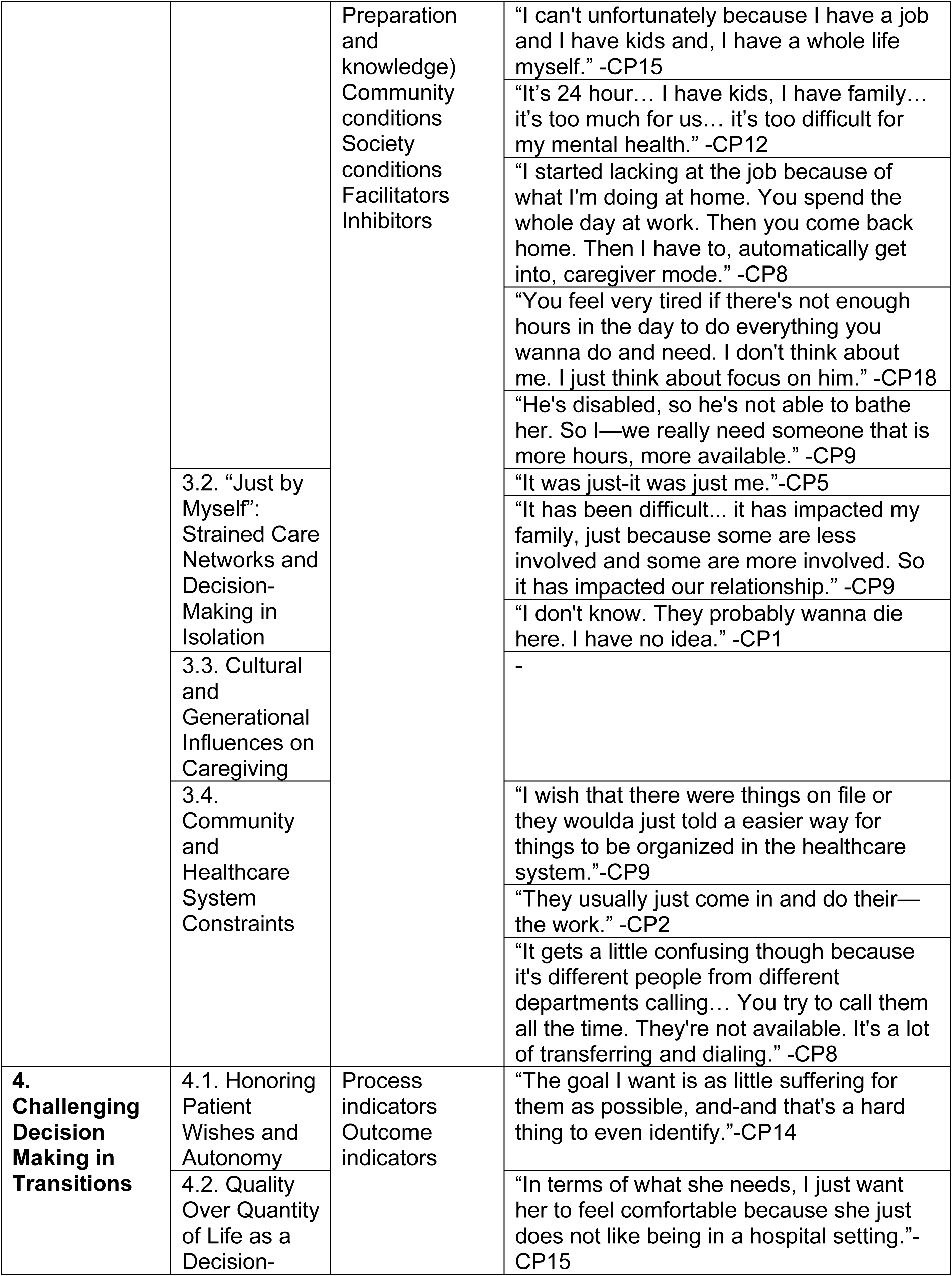
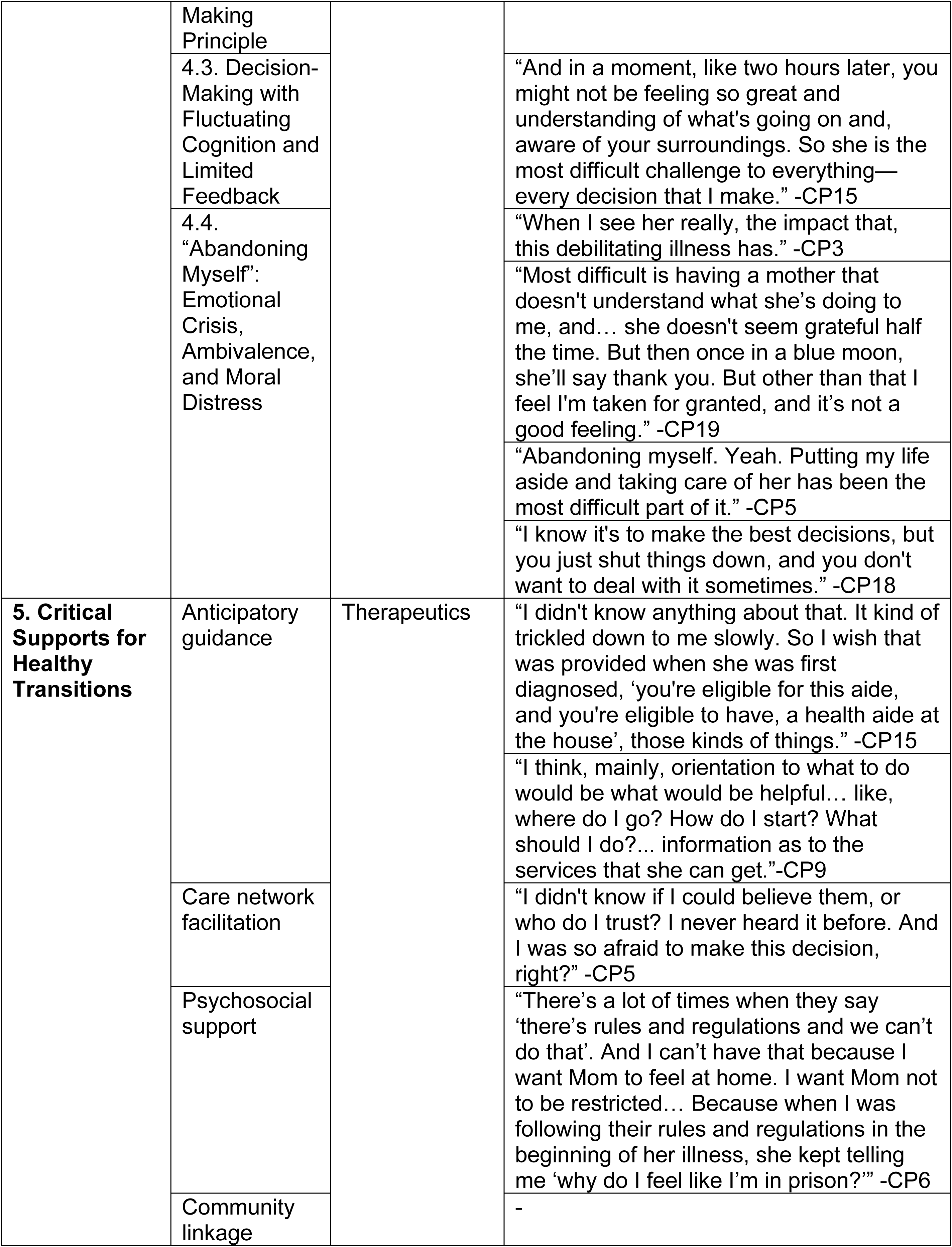
Indexing, Charting, and Interpretation.

### Findings

Participants were (Table 1) with an average age of 52.7, mostly female (80%). Participants were racially and ethnically diverse. Their educational attainment varied, from high school or GED (20%) to professional or doctoral degrees (10%). Most participants were adult children of the PWD (60%), followed by spouses (15%), grandchildren (10%), and siblings (10%). Caregiving duration was average 8.4 years. Notably, thirteen (62%) had not received information about or discussed hospice care with a healthcare professional. Participants’ care recipients PWD (n=21) had a mean age of 81.2 years and 85.7% were women. Most PWD were dual-eligible for Medicare and Medicaid (67%), duration of home care ranged from less than one year (33%) to over 20 years (5%). Diagnosis varied from Alzheimer’s disease (43%) to vascular dementia (5%). PWD had moderate to severe dementia corresponding to a mean QDRS score of 20.4.

Five major themes were identified (Table 2): (1) Multiplicity and complexity of transitions; (2) Transitions occur in unanticipated ways; (3) Challenging transition conditions, (4) Challenging decision making in transitions; and (5) Critical supports for healthy transitions.

### Theme 1: “They change and you taking the role now of the parent”: Multiplicity and complexity of transitions in dementia

Transitions in dementia are often multiple and simultaneous, affecting both PWD and their care partners. Many transitions fall into distinct but interconnected categories: developmental, situational, health-illness, and organizational. These transitions often occur concurrently and can influence one another, highlighting the complexity of dementia care.

**Developmental transitions** occur for both PWD and care partners. Aging itself is a developmental transition experienced by the older adult with dementia, while care partners undergo a significant role shift, taking responsibility for someone who may have once cared for them. For example, care partners expressed:

> *“We all have to get old. We all going to need someone to take care of us.” -CP3*
>
> *“You f-f-feel love to do it for that person, to take care of them because they’ve cared for you.” -CP4*

Some care partners spent critical amount of time parenting, waiting for years looking forward to retirement, only to face the reality that they’ve missed the opportunity to have a pre-imagined, content retirement.

> *“I thought raising kids was a thankless job but doing this was-is really-because this is supposed to be my retirement years. This is supposed to be where—you know…” -CP19*

This developmental dynamic can create a double caregiving burden in care partners of the younger generation, as many of them also care for younger family members, either children, or their siblings.

> *“It was a big struggle in the beginning for me to sort of balance her care because I have my own three children…” -CP15*
>
> *“Recently, because of my pregnancy, now, my brother’s more involved.” -CP9*

**Situational transitions** arise from changes in daily life, social engagement, and relational dynamics. PWD may experience social isolation as aging and illness limit interaction with peers, while care partners constantly adapt to new responsibilities and routines. One participant described the effects of isolation and family dynamics:

> *“I found that she was becoming—I guess we’re boring, which was really sad, at home. I guess, she felt isolated. You know, with me and the kids are young, so in her own age group.” -CP21*

Situational transitions are often related to developmental transitions. That is, under transitions of aging, people are more likely to lose loved ones around them. This loss of significant ones on PWD side risks altering their social engagement, on care partners side risks taking away their supporting network in caregiving.

> *“I lost my aunt during that time, which is my mom’s only sibling. It’s just her and her sister. And then, a year later, I lost my brother-in-law, and then—” -CP3*

A dementia diagnosis itself can be an overwhelming **health-illness transition** that equivalently has a tremendous impact on care partners:

> *“Once she told me she was sick, I burst into tears because I didn’t understand.”-CP3*

Comorbidities among PWD introduce additional layers of complexity to care transitions, as coexisting conditions can compound cognitive and functional decline and care needs. For example, one caregiver described how multiple chronic conditions intersected:

> *“His feet were very weak because he has diabetes, and he was a cardiac—open-heart patient, too.” -CP18*

Superimposed comorbidities or multiple chronic conditions of the care partner result in the individual having to navigate their own illness management and simultaneously with health-illness transitions of the PWD. A care partner expressed the challenge of caregiving while experiencing their own illness:

> *“I have sickle cell as well, so I’m not the healthiest person, but I’m trying to take care of her as best as I can, the diaper changes, the regimen, the insulin, the inhalers.” -CP21*

Due to ongoing alterations in functioning and personality, care partners experienced the gradual fading of the person they once knew. These uncontrollable changes unintentionally diminished the roles the PWD previously held in the care partner’s life. This experience represents primarily a health-illness transition, as the changes are driven by illness-related decline, and secondarily a situational transition, as the roles and relationships between the PWD and the care partner evolve.

> *“But when it comes to someone that you love and that you, see as a hero, and they change and you taking the role now kind of the parent, it’s different.” -CP7*

**Organizational transitions** occur when PWD move across care settings, such as hospitals, nursing homes, or adult day programs. These transitions often require care partners to coordinate complex logistics and advocate for the PWD:

> *“There’s a lotta forms to be filled out, a lot of, interviews and questions, it was really difficult. It was really, really hard. I don’t really know how I did it.” -CP5*

Decisions about care location also represent situational transitions, as described by a care partner reflecting on a return home from the hospital:

> *“Making decision to take her home is the best decision because after she coming back home, she totally change. Like, how she is now she can walk. In the hospital, she lay down, and she forget to open her eyes too.” -CP12*

These findings demonstrate that transitions in dementia are rarely single. Developmental changes, health decline, situational challenges, and organizational shifts often interact, creating layered and ongoing adaptation demands for both PWD and care partners. While some transitions may appear sequential such as a hospital stay leading to a nursing home placement, while the experiences are often simultaneous and related, with one transition triggering or influencing another. For example, aging and social isolation may precipitate a care role shift, while health decline can prompt hospitalization and subsequent organizational transitions.

### Theme 2: Transitions occur in unanticipated ways

Transitions in dementia rarely unfold as discrete or predictable events. Instead, they emerge through a combination of care partners’ efforts to anticipate change and the inherently unstable trajectory of the illness. Even when care partners demonstrate awareness and engagement, transitions often occur in ways that are unexpected, rapid, or difficult to interpret. Two subthemes around the following questions capture this tension: (1) how care partners orient themselves toward potential transitions, and (2) how transitions are experienced through daily fluctuations and critical events.

### Subtheme 1: Awareness and engagement shape care partners’ approach to transitions

Care partners vary in their awareness of transitions and in how actively they engage in planning and decision-making. Some demonstrate a proactive orientation, seeking information, coordinating resources, and involving family members in shared decisions regarding the care of the PWD. Decision-making is in some cases described as collective and relational:

> *“It becomes easy for me to sort of find trusted sources to then be able to think through and then make a decision. And to be clear on the decision, I have three siblings, and it’s a collaborative process where we all have different parts of what we’re doing for our family.” -CP14*
>
> *“I advocate very strongly for her… it would be a family decision. I would bring all of them in to—because I would like it to be a collective thing where we all agree together.” -CP3*

Beyond formal decisions, care partners describe an ongoing process of learning and recalibration as they encounter new aspects of the PWD’s condition. This learning often involves reassessing how they communicate, interpret behavior, and relate to the person over time:

> *“I’m talking to her like it’s normal because she has good days. When I only know how to deal with her as I’ve always dealt with her, I have to sort of readjust my lens a little bit… I’m also learning how to speak to her and navigate, this whole other side of her now.” - CP15*

Care partners also emphasize the difficulty of orienting themselves to an illness trajectory that lacks clear markers or endpoints. Unlike other caregiving experiences perceived as time-limited or developmentally bounded, dementia introduces prolonged uncertainty:

> *“The difference with an infant is your time period of doing this…With my father, I don’t know when that’s going—there’s no point where you say “they’re self-sufficient,” right?” - CP14*

These highlight the active role care partners take in anticipating transitions, while also reflecting the inherent uncertainty that shapes how transitions unfold. Notably, even among care partners who reported familiarity with dementia and confidence navigating healthcare systems, engagement was sometimes accompanied by emotional distancing. For some, managing transitions required suppressing emotional responses in order to sustain caregiving responsibilities:

> *“Working every day. It’s like, a job. I don’t think about it, to be honest. It’s like I just suppress all that.” -CP18*

### Subtheme 2: “They wake up one day, and they’re robbed of all of that”: Transitions are marked by unpredictable daily changes and critical events

Even when care partners are familiar with the PWD’s condition, dementia is characterized by constant fluctuation, including changes in cognition, behavior, mood, and physical health. Care partners described the daily unpredictability as challenging, making it difficult to anticipate the timing or impact of transitions. Daily unpredictability was described as emotionally taxing, particularly when behaviors appeared contradictory or difficult to interpret:

> *“Because her brain doesn’t work, she can’t explain herself. Then every hour, she changing. I can’t explain to you. This is too difficult and challenging.” -CP12*
>
> *“They change too much. They lose weight. They don’t want to eat. They’re crying. They’re depressed… you gotta be careful. His pressure is very bad.” -CP10*

In addition to ongoing fluctuations, care partners described critical events such as sudden functional decline, loss of communication, or rapid health deterioration. These marked significant transitions. These moments were often experienced as abrupt and destabilizing:

> *“She was eating, like a week before. I understand, dementia and all that, it can progress so, quickly, like, muscle control … But it felt a little pushed.” -CP4*
>
> *“It happened so fast that she wasn’t able to consent that I feel like I didn’t even have much time to become the caregiver, officially.” -CP9*

These experiences illustrate that transitions in dementia are not singular events but emerge through the accumulation of daily changes and situational disruptions. Care partners described a constant need to adapt as new challenges replaced those that had just been resolved:

> *“There are lots of little things that come up on a regular basis… and it’s continuous. We find that we resolve one problem, and then up comes another.” -CP11*

Transitions related to end-of-life were similarly unpredictable. Care partners frequently relied on prior experiences or observations of others to interpret signs of decline, underscoring the absence of clear guidance:

> *“I’ve learned, then, one of my neighbor, her dad, he got that problem.” -CP10*

Transitions in dementia are shaped by a tension between care partners’ efforts to anticipate change and the inherently unstable progression of the illness. While awareness and engagement influence how care partners orient themselves to potential transitions, they do not mitigate the unpredictability of daily fluctuations or critical events. These findings highlight the need for anticipatory guidance that acknowledges uncertainty, supports decision-making, and prepares care partners for transitions that unfold progressively, suddenly, and often simultaneously.

### Theme 3: Challenging transition conditions

Transitions in dementia are shaped not only by the nature of the illness but also by personal, familial, and societal conditions that can complicate the experience for both PWD and their care partners. These conditions influence care partners’ ability to anticipate, prepare for, and navigate transitions, often intensifying burden and moral distress. Three interrelated subthemes were generated around: (1) personal and caregiving burden conditions, (2) strained or absent care networks, and (3) community and healthcare system constraints.

### Subtheme 1: Personal conditions and the weight of caregiving

Personal conditions including care partner health, life circumstances, employment, family responsibilities, and the functional and medical complexity of the PWD strongly shaped transition experiences. Many care partners described significant difficulty balancing caregiving with work, parenting, pregnancy, and financial responsibilities. These competing demands contributed to exhaustion, stress, and emotional strain:

> *“It was a big struggle in the beginning for me to balance her care because I have my own three children, and she is not physically that close to me, where I could travel easily to her. So I had a lot of time constraints, and I didn’t know what was available to me.” - CP15*
>
> *“And it’s really, really difficult to hold down a job and do all this because it is so, so time consuming and, difficult to make these decisions.” -CP5*

For some, caregiving demands resulted in profound personal loss, including housing and employment:

> *“You know, I used to own a home. It’s gone. [Sniffling] I used to have a really great job. It’s gone. [Sobbing] so, yeah, the caregivers—it’s difficult.” -CP5*

Caregiving burden was further intensified by the progressive and unpredictable nature of dementia itself, particularly when PWD struggled to recognize limitations or accept care. Resistance to care and lack of insight created additional stress during transitions:

> *“Because in her mind, she can still do everything. She never was fully accepting of the fact that she would be forgetful, or maybe she didn’t want to admit it.” -CP15*

Comorbidities and complex care routines further compounded this burden, increasing both physical labor and emotional strain:

> *“She takes like 15 different medications. And then, on top of that, the therapies, and all the groceries I have to buy, the laundry is intense. There’s a lot of laundry just because she’s incontinent. I made a room for her in the kitchen. When she has a bowel movement, you can’t even go downstairs. So it’s just so challenging.” -CP21*

### Subtheme 2: “Just by myself”: Strained care networks and decision-making in isolation

Care partner well-being and access to close personal support found as critical factors. Those with engaged spouses, family members, or trusted confidants described greater capacity to manage transitions:

> *“I’m very happy that I have a caring and concerned wife, as well as cousins. It’s as a result of my wife why a lot of the issues are accomplished in an easier manner. I have someone to directly confide in. If I wasn’t married and single, oh my gosh, then I’d be in big trouble.” -CP11*

Transitions were especially challenging when care partners lacked consistent support from family or broader care networks. Many described fragmented family involvement, conflict among siblings, or complete absence of shared responsibility:

> *“No-no system at all. Just by myself, mm.” -CP2*
>
> *“I have a younger sibling and a older brother, and they just want no part of any of this as far as taking care of my mother…And it’s gotten to the point, as a family we don’t even discuss anything. Actually, we’re estranged. We don’t even talk…” -CP19*

Even when family members were present, differences in values, priorities, or interpretations of the PWD’s wishes often complicated coordination and decision-making:

> *“I became the healthcare proxy because my sisters were not in in according to an agreement with what she wanted, what she wished.” -CP3*
>
> *“Challenges are that I’ve got three other siblings, and we all don’t’ think exactly alike, right? So you’ve got to manage that component of things.” -CP14*

Without network support, decision-making transitions particularly those rooted in the PWD’s past choices or lack of advance planning, were described as emotionally overwhelming. Care partners often felt solely responsible for filling gaps left by prior decisions of PWD:

> *“Because of the decisions he made or didn’t make, now I have to step in to fill that gap for him, you know? ’Beause everything is dependent on me. That mental capacity. I’m trying my best to stay strong. And then when it comes to support, I don’t really have support really. I’m like the only daughter who’s actively doing everything.” -CP8*

When unpredictable transitions occurred in the absence of support, care partners described profound loneliness and uncertainty. They are left no choices but to make decisions in situations with little prior preference or guidance:

> *“So it’s a very lonely place to make all of the decisions on my own. It really is. And sometimes I-am I doing the right thing?” -CP19*

### Subtheme 3: Cultural and generational influences on caregiving

Care transitions in dementia are shaped also by cultural norms, generational differences, and moral expectations within families. These factors can complicate communication, decision-making, and coordination of care. Care partners described challenges navigating differences in worldview or caregiving philosophy across generations:

> *“My mother is still of the very traditional world. One major challenge has always been this inability to articulate ourselves and understand the subtleties of what she was trying to say… It’s the reasoning for the care to be explained to someone of a different generation and of a different culture.” -CP14*

Social and moral pressures within communities and families also intensified caregiving burden, particularly when decisions such as nursing home placement were judged or challenged by others:

> *“My cousin put his mom, my aunt, in a nursing home. She was lighting fires. He and his wife were both working and couldn’t keep an eye on her. Everyone stopped talking to him. People fought with him all the time. Other family members tried to break her free from the nursing home. It was total drama.” -CP21*

In some cultural contexts, care responsibilities are seen as obligatory, which can create tension when the care partner is unwilling, unable, or constrained by other responsibilities:

> *“In Muslim culture, parent should be the kid’s responsibility… but she is mom, and my husband doesn’t like—he can’t change the diaper.” -CP12*

This subtheme highlights that dementia care is embedded in broader social and cultural frameworks. Cultural and generational expectations can amplify caregiver stress, complicate care planning, and shape how transitions are experienced and negotiated.

### Subtheme 4: Community and healthcare system constraints

Broader community and healthcare system factors also played a critical role in shaping transition experiences. Care partners described system-level barriers including rigid eligibility criteria, fragmented services, administrative burden, and understaffing, which often delayed care and complicated the transitions.

> *“That was also difficult because you need to meet certain criteria, which she never met during the interviews because she would just completely deny her situation.” -CP15*
>
> *“It is overwhelming. It was a lot. It was just the sheer volume of the phone calls, the physical therapy, the occupational therapy, the speech therapy, the nursing calls, the peace manager from the nursing agency, and then the agencies themselves, so much issues with them.” -CP21*

Administrative inefficiencies and insurance barriers were described as especially taxing during transitions:

> *“What kills us is really the administrative issues that go behind. For example dealing with Medicaid, dealing with Medicare, dealing with the specific procedures for the doctors, for example” -CP14*
>
> *“You have to jump through all these hoops to get one thing and then make phone calls, and then the doctor has to sign off on it. And by the time that happens, it’s like a week or two later. And so, I was getting calls like everyday for my mom, stressing out.” -CP19*

Care partners frequently expressed frustration with having to navigate complex systems without adequate guidance, particularly following diagnosis:

> *“I guess the healthcare system, I wish that it was more understandi—or it would be a easier transition, like as soon as she had the diagnosis, that at least, someone woulda spoken with me, let’s say, at her general medical provider’s office, and givin’ me the paperworks needed where I can just have authorization to create an appointment” -CP9*

Staffing shortages further undermined confidence in care settings. Care partners consistently noted staffing limitations and time pressures in care settings, which can compromise care quality and create stress during transitions:

> *“There are fabulous nurses and great staff, they’re just understaffed. Some patients suffer a little, and I don’t want that for my mom.” -CP21*

These challenges were often compounded by how healthcare professionals communicated and assisted decision-making during care transitions. Care partners frequently perceived healthcare interactions as protocol-driven and task-oriented, with limited attention to the relational knowledge care partners held about the PWD or to the broader dynamics within caregiving dyads:

> *“Be reasonable about how they help her and not just, “Well, I’m not allowed to do this, but I am allowed to do this,” that kind of stuff.” -CP15*

This approach was particularly challenging during organizational transitions, such as hospitalization or transfer to long-term care settings. Care partners emphasized that such transitions disrupted familiarity and continuity, which are critical for PWD, yet were often treated as routine or unavoidable within the healthcare system:

> *“When she hospitalized, a doctor told me we have to send her to the nursing home. But the thing is, because she have dementia, she only know me. Because I’m taking care of her and living with her, she only know my name, not my face.” -CP12*

Despite these challenges, care partners also identified conditions that facilitated smoother transitions, including continuity of care, reliable insurance, and accessible home- and community-based services:

> *“He’s been seeing the same people, so they have a record of everything now with computer. They can just pull everything up and look there.” -CP18*
>
> *“Thank God for the insurance… the homecare agency calls it keeping you out of nursing care at home. It’s 24-hour care. We coordinate with family members and aides. It’s been very good, working very well for her.” -CP21*

Day programs and home care services were particularly valued for improving PWD well-being and providing respite for care partners:

> *“The aide mentioned senior center. And wow, that is great… she comes back from that place a different person, and she’s so content. She’s smiling.” -CP21*

### Theme 4: Challenging decision making in transitions

#### Subtheme 1: Honoring patient wishes and autonomy

Care partners described their responses to transitions in dementia as a continuous process of prioritizing quality of life of PWD. Care partners frequently referenced previously expressed wishes of the PWD as a guide for decision-making. PWD’s wishes served as anchors during periods of uncertainty:

> *“I just wanna honor whatever she ask because it’s not about me. It’s about her, you know?” -CP3*
>
> *“Sometimes actually asking her… that person should still have a say, even if you think they’re not gonna, even what they’re having a say on perhaps. So sometimes still kinda asking her.” -CP4*

*Decision-making extended to micro-level daily choices throughout the dementia trajectory*:

> *“I would like to see what would be on the menu for her. She always liked magazines. So I would ask if that be available for her. For her, they’re comforting. She likes to watch Spanish shows. So I would prefer that she would have a television because that’s how her day goes.” -CP6*

Respecting autonomy was not a one-time decision, but an ongoing interpretive process in dementia, requiring care partners to translate prior wishes into present-day choices amid changing clinical realities. Care partners described drawing on intimate knowledge of PWD’s routines, preferences, and personality to guide these everyday decisions, often using small environmental or activity-related adjustments to preserve familiarity and comfort. In this way, autonomy was enacted not only through formal care decisions but also through efforts to maintain continuity in the person’s daily experiences and sense of self.

#### Subtheme 2: Quality over quantity of life as a decision-making principle

Many care partners articulated a clear prioritization of quality over quantity of life, particularly when faced with decisions about hospitalization, aggressive treatment, or institutional placement. This orientation is often the coping strategy to navigate morally complex decisions:

> *“The quality of life is gone and there’s nothing there. At that point, truthfully, I would not want to keep her hanging on.” -CP3*

Home-based care was frequently described as a means of preserving dignity, comfort, and function, in contrast to perceived decline in institutional settings:

> *“I want her to be in the place that she’s most comfortable and familiar with.” -CP3*

These experiences reinforced care partners’ commitment to avoiding organizational transitions when possible. This quality over quantity principle shaped care partners’ willingness to accept trade-offs.

#### Subtheme 3: Decision-making with fluctuating cognition and limited feedback

Care partners frequently expressed the weight of making decisions for another person, often without feedback from the PWD or the healthcare system:

> *“Just, sometime, making decisions and knowing that I’m making the right decision when she can’t make the decision. Oh God, if I get it and something happens, you know, oh, the blood is gonna be on my hands. And so I had a lotta mix emotions about that.” -CP3*

Care partners relied on research, professional guidance, and prior knowledge of the PWD’s preferences, while continued to doubt whether their decisions were optimal, especially as cognition fluctuated.

> *“The most challenging is having to, think for someone else… am I making the best decision? … I’m basing it off, research sometimes, or, what the doctor’s recommending. So it’s always, is it the best decision? That’s the biggest difficulty.” -CP8*

#### Subtheme 4: “Abandoning myself”: Emotional crisis, ambivalence, and moral distress

The weight of decision-making, coupled with caregiving demands, contributed to emotional crises, ambivalence, and moral distress. Care partners described feelings of love and devotion coexisting with exhaustion, guilt, frustration, and sometimes wishing for an end to suffering.

Doubt rooted in decision making often shifted from “am I making the best decision” to “am I doing the best I can”.

> *“Sometimes it gets overwhelming… I get frustrated. I feel like I’m not doing the best that I can.” – CP4*

As close observers of their loved ones’ decline, care partners struggled with the emotional toll of witnessing the illness’s impact:

> *“It’s very hard to see your loved one decline irregardless of what the decline is, whether it be, mental or physical.” -CP7*

Emotional ambivalence was common, with care partners balancing love and devotion with feelings of being unappreciated or overwhelmed:

> *“I always say to myself, she doesn’t do this with intent or to drive me crazy or to have me pull my hair out.” -CP7*

In the most emotionally intense accounts, care partners described reaching a breaking point:

> *“I don’t have the time, and I don’t have the mental energy…your mental health gets so affected by taking care of somebody. Caregivers just don’t have enough support… I wish she’d, like, leave. I wish she crossed over because I couldn’t do it [crying].” -CP5*

These narratives reflect moral distress, where care partners struggle to reconcile deeply held values of love, duty, cultural expectations, and respect for autonomy, with the relentless demands of caregiving.

Across these four themes, this study revealed that transitions are multiple, simultaneous, and related. Transitions in dementia are unpredictable and shaped by personal and systemic conditions. Responses to transitions are guided by an enduring commitment to autonomy and quality of life of PWD. Theme 1 highlighted the complexity of simultaneous developmental, health-illness, situational, and organizational transitions. Theme 2 underscored how variable awareness and unanticipated changes challenge care partners’ ability to anticipate and navigate transitions. Theme 3 illuminated the personal, cultural, and systemic conditions that exacerbate stress and moral distress. Theme 4 demonstrated how care partners respond by prioritizing the PWD’s preferences, navigating decision-making difficulty, often under considerable emotional strain.

### Theme 5: Critical supports for healthy transitions

Care partners’ experiences highlight the urgent need for structured transitional care support consistent with the concept of nursing therapeutics, in which healthcare professionals assess readiness, prepare individuals and families for change, and supplement caregiving roles through coordination and guidance [11]. Interventions in this context facilitate transitions by providing anticipatory guidance, mediating care networks, providing psychosocial support, and linking PWD and care partners to available resources.

Care partners frequently described feeling under-informed, uncertain, and unsupported in navigating the complex landscape of dementia care:

> *“[Sighs] I don’t think I know as much as I should… fearful that they might not get the care that they need … you’re wracking your brain because you wanna make the best decision for your loved one, and you’re not informed.” -CP5*

Care partners emphasized the desire for comprehensive knowledge and options to guide decision-making:

> *“So having those options and just not one option presented. Having whatever options can be available.” -CP4*

Most care partners described independently gathering information but expressed a preference for guidance from trusted, authorized sources:

> *“The education is most important because they know more than I do. There’s so much I can research, but they-they’ve experienced it.” -CP8*
>
> *“You’re in such an emotional place, and you’re sleep deprived, and your brain can only handle so much information in one day. So I think having one contact person or two contact people and having it written out for you…” -CP5*

Care partners described tangible benefits when PWD were connected to geriatric practices, social workers, senior centers, and educational workshops, which enhanced functional and emotional well-being for PWD while also strengthening care partners’ preparedness and confidence in decision-making.

> *“Information or workshops about how this all works, would be helpful too, like forums, Zoom things that people could go on and learn, what is an MLTC? What is a waiver program? How do you pick a home health agency?… I think caregiver-givers need somebody to talk to… they just need somebody to hear what they’re going through… which I’m sure it’s out there, but, how do you find this stuff when you’re so busy taking care of your loved one?” -CP5*

Importantly, participants emphasized that guidance and interventions should prioritize daily preferences, routines, and personal narratives that bring comfort and continuity to both PWD and care partners. Transitional support should allow flexibility, serving as a guide rather than rigid rules, and adapt to the unique values and cultural practices of each family. While standards and best-practice recommendations can inform care, they should not override approaches that preserve familiarities that support well-being.

> *“If the person—if they’re able to share their wants and needs, will that be met? Strictly by doctor’s orders or whatever it is?… It’s difficult sometimes, because you’re balancing, like your religion and then what the doctor’s saying sometimes. So it can be tricky.” -CP8*

Care partners also stressed the need for resource linkage and a shared information system for programs such as daycare, respite care, and community services. They highlighted challenges related to inconsistent information and trust:

> *“My doctor, and I’m go tell them about it, too, he’s like, “I don’t think there’s any day programs she would be able to go to.” -CP7*

These findings suggest that transitional care can operationalize by integrating anticipatory guidance, care coordination, psychosocial support, and community linkage. Thereby mitigating the complexity and unpredictability of transitions in dementia. As indicated in Table 3, transitional care should facilitate anticipatory guidance to prepare PWD and care partners for transitions, honor routines and preferences while maintaining safe care practices, iInclude psychosocial support for care partners to enhance adaptation, and coordinate resources and establish a central contact to foster trust and reduce burden.

**Table 3.** Transitional Care Recommendations.

|  |  |
| --- | --- |
| **Anticipatory Guidance** | Facilitate anticipatory guidance to prepare PWD and care partners for transitions |
| **Care Coordination** | Honor routines and preferences while maintaining safe care practices |
| **Psychosocial Support** | Include psychosocial support for care partners to enhance adaptation |
| **Community Linkage** | Coordinate resources and establish a central contact to foster trust and reduce burden |

### Implications

Applying Transitions Theory allowed this study to capture a breadth of these transition experiences. In return, this study tested and refined the conceptual model as indicated in Fig 1. Transitions in dementia are often unanticipated and continuous, where decision-making is a core property. Care networks, community resources, cultural and generational factors, logistics navigating care were critical systemic conditions for transitions. Several process and outcome indicators including autonomy, quality of life, decision-making, psychosocial well-being, cognitive and functional capacity were articulated.

**Fig 1.**
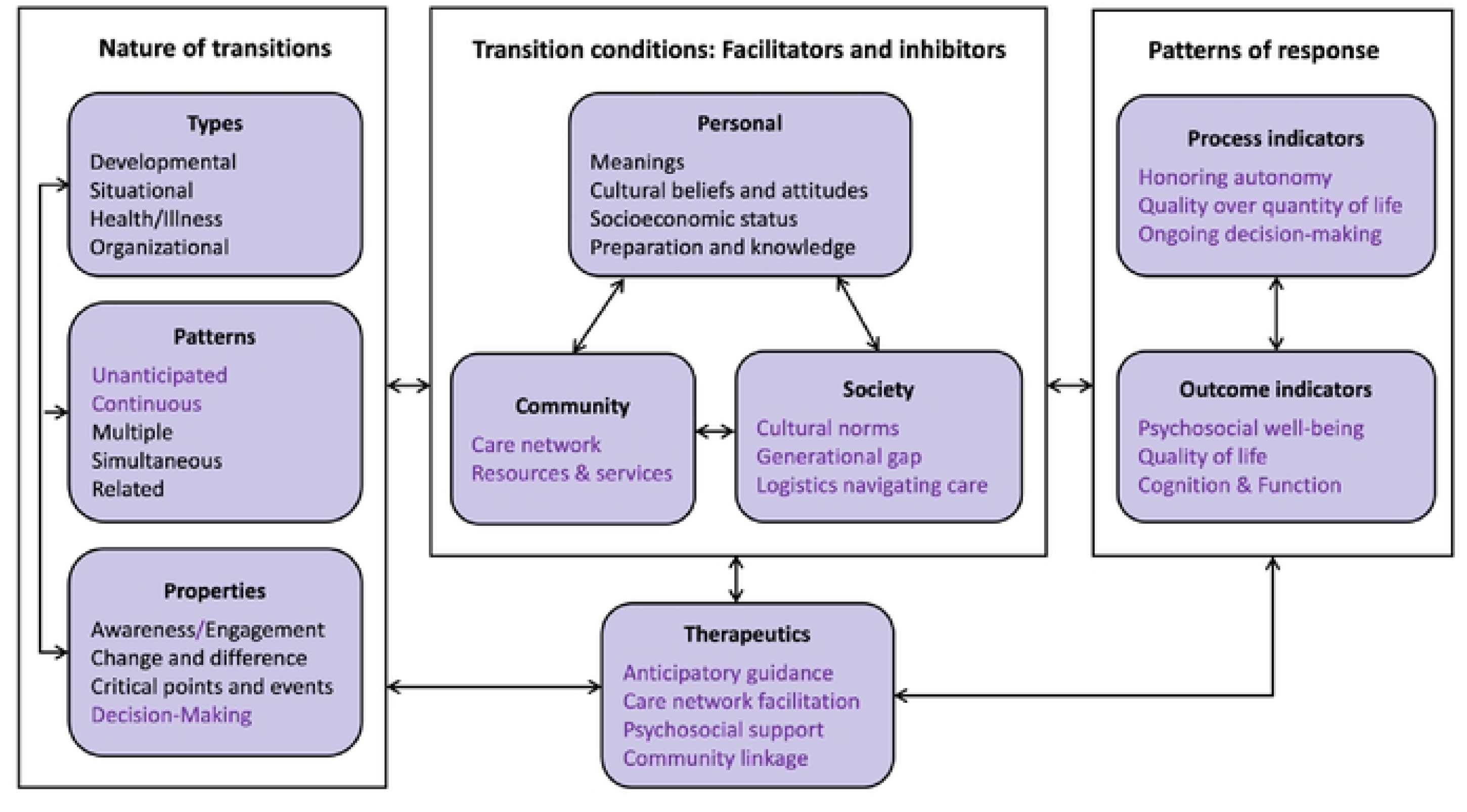
Transitions in Dementia Framework, Adapted from Transitions Theory. The framework illustrates key transition processes across the dementia trajectory. Colored components represent elements that were added or modified from the original Transitions Theory framework to address dementia-specific contexts and experiences.

This study explored the transitional care needs of PWD and their care partners applying a secondary qualitative analysis. The findings highlight that transitions in dementia are complex and often unanticipated, involving simultaneous developmental, health-illness, situational, and organizational changes. PWD experience developmental transitions associated with aging and health-illness transitions related to dementia, care partners undergo developmental transitions marked by shifts in caregiving roles. Both groups continuously experience situational transitions due to the unpredictable trajectory of dementia particularly in later stages, organizational transitions as care moves across settings. These transitions are closely interrelated. For example, aging may contribute to situational social isolation; caregiving role transitions can necessitate organizational placement in long-term care. These findings extend previous research that underestimate the layered and overlapping transitions experienced by PWD and their families.

These highlight the multiplicity and complexity of transitions in dementia, underscoring the need for transitional care approaches that are responsive to the evolving needs of both PWD and their care partners. Care partners’ responses to dementia transitions are characterized by value-driven decision-making under sustained burden. Prioritizing quality of life functions as both a guiding principle and an adaptive response, helping care partners navigate uncertainty and moral complexity. However, the emotional toll of sustaining this orientation indicates the need of adequate support. That is transitional care approaches that not only honor patient wishes but also actively address caregiver well-being, normalize ambivalence, and provide structured guidance before and during both high- and low-stakes decision points.

The findings also demonstrate that care partners overwhelmingly rely on self-directed research due to gaps in system-provided guidance. This burden compounds the challenges of decision-making in the context of cognitive decline and functional loss in PWD. Care partners navigate transitions while balancing competing life demands and often compensating for limited or fragmented support networks. Transitions are further shaped by cultural and generational factors. Care partners described challenges navigating differences in worldview, caregiving philosophy, or moral expectations across generations, which could complicate communication, decision-making, and coordination of care.

System-level constraints further complicate transitions, not only through administrative and resource limitations, but also through interactional practices within healthcare settings. Care partners suggest that standardized, protocol-driven approaches to care can overlook their experiential knowledge and the relational importance of familiarity for PWD. Organizational transitions such as hospitalization or placement changes were especially destabilizing when decisions were made without attention to established caregiving relationships or the lived context of dementia care.

Apart from such high-stake decisions, daily micro-level decisions illustrate how care partners actively reconstructed autonomy in reality. They balance safety, clinical recommendations, and institutional constraints while attempting to honor the individual’s identity and past preferences. Over time, this interpretive role positioned care partners as key mediators between PWD’s evolving needs and the structured environments, requiring ongoing judgment, advocacy, and physical emotional labor.

Notably, these experiences and views illustrate how system-driven decision-making and standardized procedures can overlook the relational and contextual realities of dementia care. Organizational transitions, when enacted without sufficient recognition of familiarity, attachment, and caregiver knowledge, were experienced as particularly destabilizing for both PWD and care partners. Healthcare system constraints are not limited to access or resources but also include interactional practices that shape how transitions are negotiated and experienced.

These challenging transition conditions reveal that dementia-related transitions unfold within a complex context of personal circumstances, relational dynamics, cultural norms, and healthcare system structures. Together, these findings highlight that dementia care transitions are socially, culturally, and structurally situated processes. Transitions are determined by the interplay of individual care partner capacity, family and care network dynamics, generational and cultural influences, and healthcare system practices. Capturing this broader context is essential to understanding the full complexity of dementia care as it is lived by care partners and PWD.

Finally, this study underscores the critical role of healthcare professionals as facilitators of transitions, aligning with Transitions Theory. Participants’ views illustrate that interventions such as referral to geriatric practices, linking to social workers or senior centers, and providing workshops on navigating services, can reduce caregiver strain during transitions. That is, transitional care interventions that address knowledge, coordination, emotional support, and resource linkage can strengthen dementia care, promoting healthy, value-driven transitions.

### Limitations

As a secondary analysis, this study has inherent limitations. The parent study focused on hospice and end-of-life care transitions, and data were collected primarily to address those aims. Although the data were re-coded and deemed saturated for the research questions of the present analysis, this focus may have limited variation relevant to the broader goal of characterizing transitions in the full illness trajectory. The cross-sectional design further constrained the ability to examine transitions over time, observe sequential processes, or assess relationships between triggering events and subsequent transitions. Despite these limitations, this study provides in-depth insights into transitions in dementia and the transitional care needs of persons with dementia and their care partners.

Future research should focus on longitudinal exploration of dementia transitions to capture more transition-related concepts. Understanding these dynamics could inform timely, anticipatory, and scalable interventions, including structured programs or guideline-based checklists, to support healthy transitions throughout the dementia trajectory.

## Data Availability

No data was generated by this study.

## Notes

### Competing Interest Statement

The authors have declared no competing interest.

### Author Declarations

Approved by the institutional review boards of NYU Langone Medical Center and VNS Health.

